# Transmission potential of human monkeypox in mass gatherings

**DOI:** 10.1101/2022.06.21.22276684

**Authors:** Vana Sypsa, Ioannis Mameletzis, Sotirios Tsiodras

## Abstract

Since May 2022, a large number of monkeypox cases has been reported in non-endemic settings. Taking into account the strict measures implemented due to the COVID-19 pandemic and the desire of people to reclaim what is perceived as lost time, it is anticipated that mass gatherings this summer will be highly attended. Based on data for the secondary attack rate among unvaccinated contacts from endemic countries, we estimate that, on average, more than one secondary case is anticipated per infectious person if he/she has a high number of group contacts (>30) or more than eight close contacts. Although the role of group contacts in mass gatherings is uncertain (less likely to involve physical contact, shorter duration), close contacts associated with the event (e.g. intimate/sexual contact with other attendees) might be the amplifying event. Enforcing awareness, early recognition and engaging affected populations in the monkeypox response are important to control transmission.

---

Since May 2022, a large number of monkeypox cases has been reported in Europe, the US and other non-endemic settings [1]. To date, most cases have been identified, but not exclusively, in gay, bisexual and other men who have sex with men (MSM). In Europe, several of these cases are associated with saunas and fetish festivals, whereby individuals were exposed to the virus through close contact [2, 3]. It is known that social and mass gatherings constitute high risk settings for the transmission of infectious diseases due to close contact, especially in crowded venues [4]. Furthermore, attendance by people travelling from other countries can further facilitate the introduction and spread of novel pathogens. Taking into account the strict measures implemented in many countries for more than two years due to the COVID-19 pandemic and the desire of people, including young persons, to socialise, celebrate, and reclaim what is perceived as lost time, it is anticipated that these events will be highly attended. In this perspective, we assess the likelihood scenarios for monkeypox transmission in venues and settings where conditions are conducive to amplifying spread, we highlight the challenges in this endeavour and emphasise the importance of community engagement and risk communication.

## Mass gatherings and social contacts

During the summer, a large number of social/mass gatherings are commonplace, including music festivals that attract thousands of people, such as Glastonbury in the UK (approximately 200,000 people/day), the Zurich Street Parade in Switzerland (techno party with estimated 850,000 people in 2019), and Summerfest in Wisconsin (about 850,000 people attend several days) [5, 6]. After COVID-19 related restrictions imposed in the two past years leading to cancellation or to attendance restrictions, events this summer are anticipated [7]. Apart from music festivals, other events are being organised such as parties in popular travel destinations (e.g. Ibiza in Spain or Mykonos in Greece).

The number of contacts in mass gatherings cannot be easily ascertained. From social contact surveys, the mean number of contacts on a Saturday in Germany (including group contacts) was 19.5 [8], whereas the average size of group contacts in the UK was 20.3 with one fourth of them being physical [9]. However, estimates on the number, duration and type of contacts obtained from this type of surveys cannot be reliably extended to the setting of mass gatherings. Other approaches are necessary. For example, social mixing patterns at an event have been estimated using video analysis technology [10]. An experiment to measure direct contacts was performed in an indoors mass gathering (seated concert) where contact tracing devices were handed out to approximately 1100 attendees [11]. In this concert, there were contacts with on average 8.9 and 14.1 different persons lasting for more than 15 and 5 minutes, respectively. Although such data are valuable, they may not reflect the nature of contacts in social/mass gatherings described here in a context where thousands of people congregate for more than a few hours.

## Estimates of human-to-human transmissibility of monkeypox virus

The increase in the number of reported monkeypox cases in endemic countries over time, in parallel with the declining vaccination coverage for smallpox [12] and the current outbreaks in non-endemic countries confirm the transmission potential from human to human. The available data on R_0_ are limited and not contemporary [13]. In fact, estimates were derived from endemic countries in past years where a large proportion of the population was vaccinated against smallpox. When this was taken into account in a recent paper, R_0_ in a completely susceptible population was estimated to be around 2 [14]. There are, however, available estimates of the secondary attack rates (SAR) - separately for household and non-household as well as for vaccinated and unvaccinated contacts - that can be used to estimate R_0_ [15-18]. More specifically, if transmission is stratified by contacts within and outside of the household, then R_0_=SAR_1_·N_1_+ SAR_2_·N_2_ (equation 1), where SAR_1_, SAR_2_ are the secondary attack rates and N_1_, N_2_ are the numbers of at-risk contacts made within household and wider community, respectively [19, 20].

## R_0_ for monkeypox in the setting of social/mass gatherings

We attempted to explore the transmission potential of human monkeypox in mass gatherings under various scenarios for the number of close and other (group) contacts. We assumed that an infectious person attending such an event will have 2-10 close contacts (e.g. family, friends) as well as contacts with other attendees (ranging from 5-50). We obtained R_0_ using equation 1 where we applied available estimates of the secondary attack rate for unvaccinated contacts [15]. More specifically, we used the household and non-household SAR estimate for close contacts and for contacts with other attendees, respectively. To account for the uncertainty in SAR, we performed simulations where we obtained SAR from a normal distribution using the estimated mean and 95% CI from a meta-analysis (estimate [95% CI]: 7.57 [0, 15.17] for unvaccinated close contacts and 2.69 [0, 6.17] for unvaccinated other contacts)[15]. For each scenario for the number of contacts, the mean R_0_ and 2.5th and 97.5th percentiles were obtained through 5,000 simulations.

Based on the simulations (Figure 1), the mean R_0_ could reach up to 2.1 in the case of a high number of close and other contacts. R_0_ could exceed 1 if the infectious person has contacts with more than approximately 30 persons even if he/she has close contacts with only 2-3 persons. Similarly, a high number of close contacts (8-10 contacts) could lead to more than one secondary case even if the number of other contacts is relatively small (less than 15).

**Figure 1.**
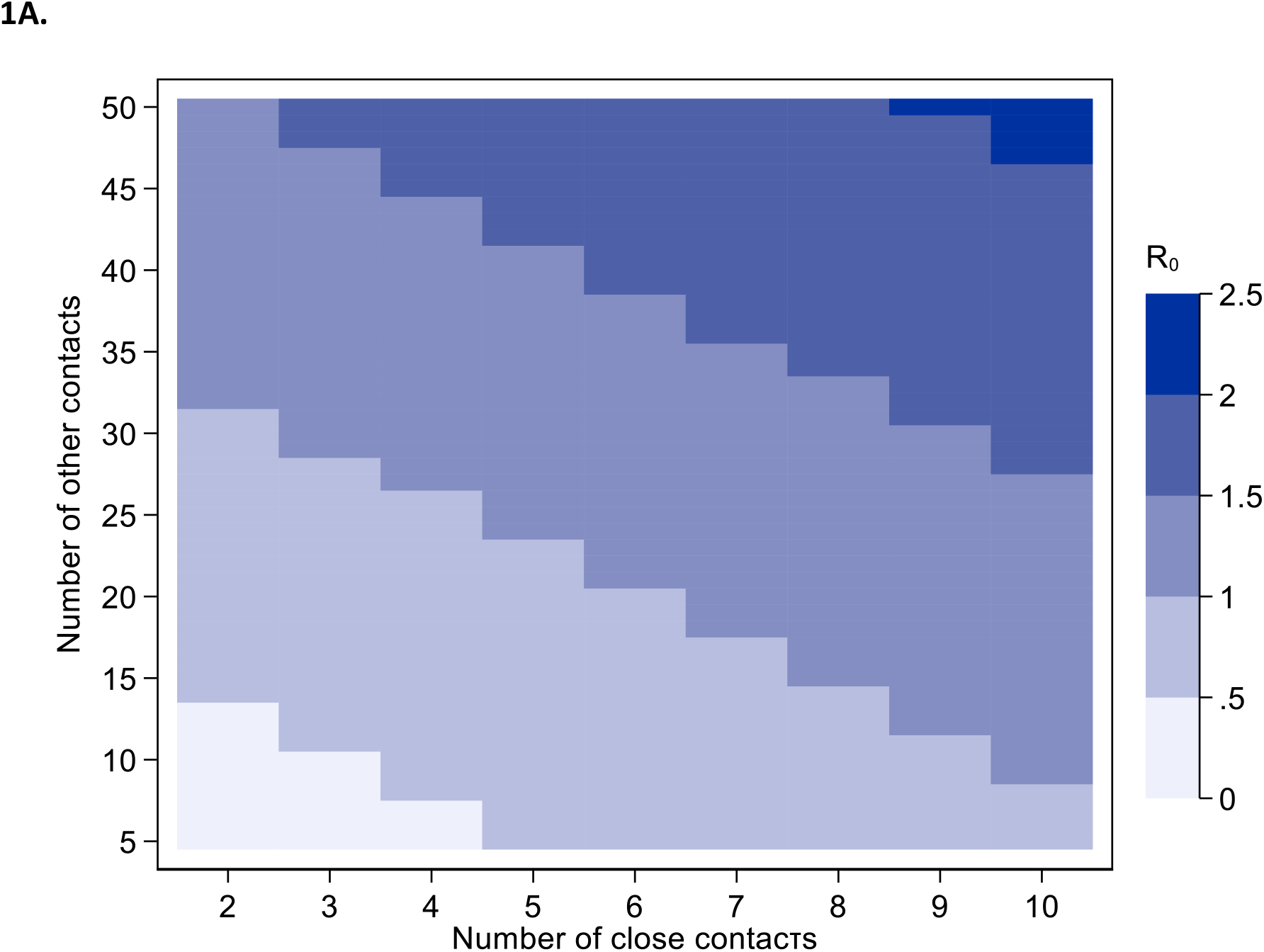
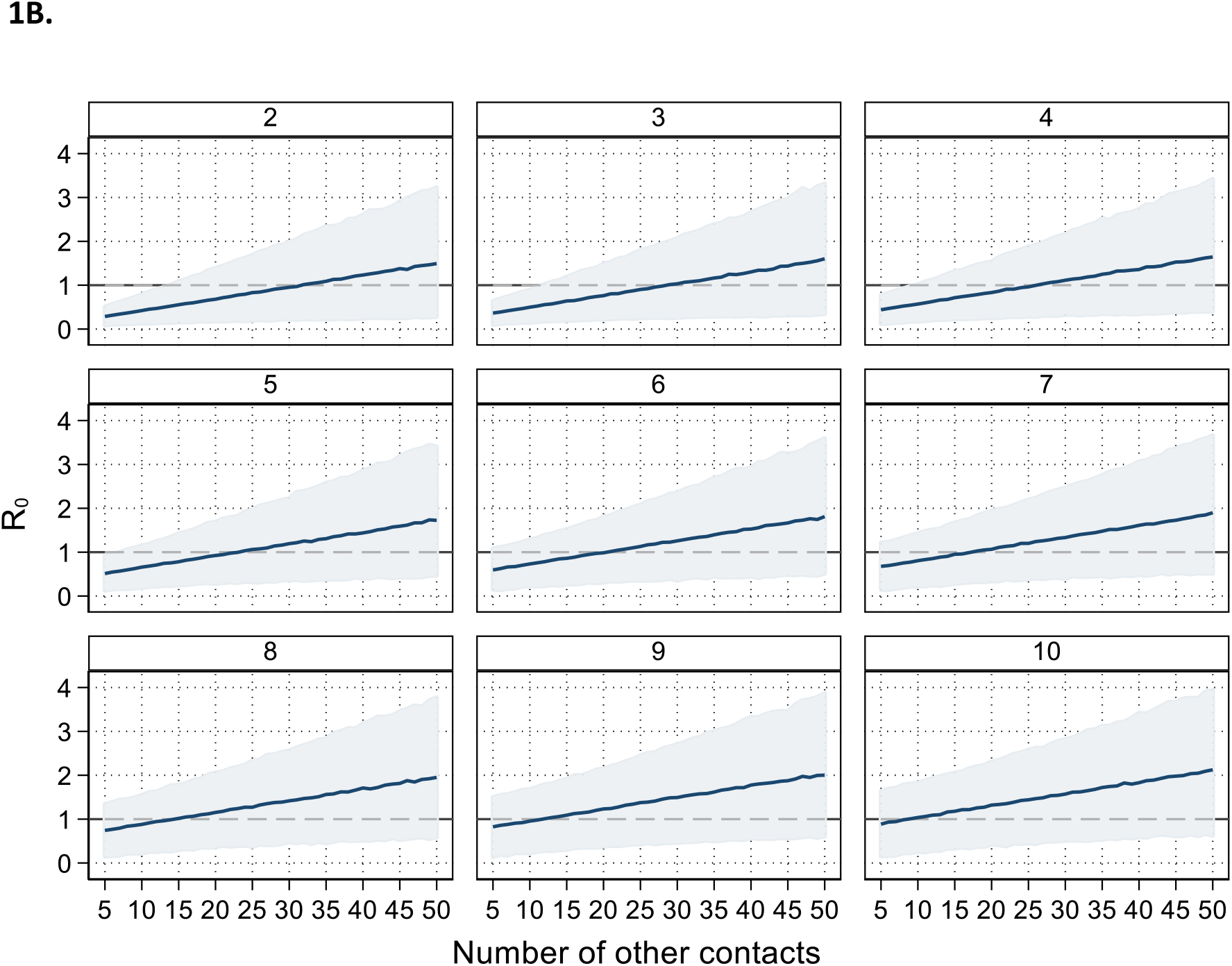
Basic reproduction number (R_0_) estimated under various scenarios for the number of close and other (group) contacts in a mass gathering. Mean and 95% credible intervals were obtained through 5,000 simulations with secondary attack rates for unvaccinated contacts obtained from[15]. **A**. Heatmap of the mean R_0_ **B**. Mean (thick lines) and 95% credible intervals (shaded areas) of R_0_. The dashed line indicates the threshold of R_0_ = 1. Sub-graphs correspond to the respective number of close contacts.

## Discussion

There is a potential for increased transmission of human monkeypox in non-endemic areas during social/mass gathering in the following months. However, our estimates should be interpreted with caution taking into account several limitations. First, we assumed a totally susceptible population given that smallpox vaccination ended in early 1970s in Europe and the US and that mass gatherings, including concerts, are mainly attended by younger people. Second, the secondary attack rates we have used were obtained from data collected several years ago in endemic countries; other factors may have contributed to increased transmission in these settings besides lack of immunity like poor hygiene and crowded living quarters. Third, the secondary attack rate is the probability that a case infects a contact over his or her infectious period whereas mass gatherings may have a duration of a few hours up to a few days. However, the SAR estimate for non-household contacts that we have used to assess transmission among non-close contacts in a mass-gathering may be still appropriate as non-household contacts do not last over the whole infectious period and could even be one-time event.

The number of effective contacts, i.e. contacts sufficient to lead to monkeypox transmission between an infectious and a susceptible individual in the setting of mass gatherings is uncertain. For instance, in concerts with thousands of attendees, it is anticipated that the number of contacts will be high. However, compared to individual contacts, group contacts are less likely to be physical. In addition, there is a saturation of contact duration for individuals with large numbers of contacts [9]. As a result, the role of these encounters in transmission is uncertain. It is also possible that close contacts associated with the event (but not taking place during the event, such as intimate or sexual contact with other attendees) could be the amplifying event. In our simulations, eight to ten close contacts could lead to more than one secondary case, on average, per index case even if the number of other contacts is relatively small.

## Conclusions

There is an inevitable comparison of this new outbreak in non-endemic countries to COVID-19 pandemic. Nonetheless, it is known that the success of control measures in an epidemic is determined by two factors: the proportion of transmission occurring prior to the onset of clinical symptoms and the transmissibility of the pathogen as measured by R_0_ [21]. Monkeypox is not as efficiently transmitted as SARS-CoV-2 and is considered to be mainly contagious after the development of symptoms. Thus, in view of mass gatherings and relaxation after two years of social restrictions, enforcing awareness and early recognition is important to control transmission and stop the spread. Lastly, and importantly, public health authorities, governments, and other stakeholders should make every concerted effort to ensure that no population affected by monkeypox is stigmatised. Instead, the people currently most affected in the current outbreak, MSM, should be actively engaged in the monkeypox response, including in the critical area of risk communication and community engagement, which is at the core of an effective public health response [22].

## Data Availability

Not applicable (simulations using estimates from the literature)

## Ethical statement

No ethical approval was required as the analysis was performed using estimates from the literature

## Author contributors

All authors fulfilled the ICJME authorship criteria. VS prepared the first draft and performed the simulations. ST and IM gave critical feedback and contributed to the literature search, data interpretation, writing and editing. All authors approved the final version of the manuscript.

## Conflict of interest

We declare no competing interests related to this work.

## Financial support

None

## Notes

### Competing Interest Statement

The authors have declared no competing interest.

### Funding Statement

This study did not receive any funding

